# Distinct and joint effects of low and high levels of Aβ and tau deposition on cortical thickness

**DOI:** 10.1101/2022.09.09.22279694

**Authors:** Seyed Hani Hojjati, Tracy A. Butler, Gloria C. Chiang, Christian Habeck, Arindam RoyChoudhury, Farnia Feiz, Jacob Shteingart, Siddharth Nayak, Sindy Ozoria, Antonio Fernández, Yaakov Stern, José A. Luchsinger, Davangere P. Devanand, Qolamreza R. Razlighi

## Abstract

Alzheimer’s disease (AD) is defined by the presence of Amyloid-β (Aβ), tau, and neurodegeneration (ATN framework) in the human cerebral cortex. Prior studies have suggested that Aβ deposition can be associated with both cortical thinning and thickening. These contradictory results may be due to small sample sizes, the presence versus absence of tau, and limited detectability in the earliest phase of protein deposition, which may begin in young adulthood and cannot be captured in studies enrolling only older subjects. In this study, we aimed to find the distinct and joint effects of Aβ and tau on neurodegeneration during the progression from normal to abnormal stages of pathologies that remain incompletely understood. We used ^18^F-MK6240 and ^18^F-Florbetaben/^18^F-Florbetapir positron emission tomography (PET) and magnetic resonance imaging (MRI) to quantify tau, Aβ, and cortical thickness in 529 participants ranging in age from 20 to 90. We applied a novel partial volume correction technique based on the absence of proteinopathy in young controls to optimize spatial resolution. Aβ/tau abnormality was defined at 95^th^ percentile of the normal distribution of global Ab/tau observed in young participants. We performed multiple regression analyses to assess the distinct and joint effects of Aβ and tau on cortical thickness. Using 529 participants (83 young, 394 healthy older, 52 MCI) we showed that normal levels of Aβ deposition were significantly associated with increased cortical thickness regardless of the amount of tau (e.g., left entorhinal cortex with t>3.241). The relationship between tau deposition and neurodegeneration was more complex: abnormal levels of tau were associated with cortical thinning in several regions of the brain (e.g., left entorhinal with t<-2.80 and left insula with t<-3.202), as expected based on prior neuroimaging and neuropathological studies. Surprisingly, however, normal levels of tau were found to be associated with cortical *thickening*. Moreover, at abnormal levels of Aβ and tau, the *resonance* between them, defined as their correlation throughout the cortex, was associated strongly with cortical thinning when controlling for their additive effect. We confirm prior findings of an association between Aβ deposition and cortical thickening and suggest this may also be the case in the earliest stages of deposition in normal aging. We discuss potential pathophysiologic processes underlying this effect such as inflammation and hyperactivation (excitotoxicity). We also illustrate that resonance between high levels of Aβ and tau uptake is strongly associated with cortical thinning, emphasizing the effects of Aβ/tau synergy in AD pathogenesis.

## Introduction

Alzheimer’s disease (AD) is a neurodegenerative disorder that significantly impacts the brain’s structure and function [1-3]. Amyloid-β (Aβ) and tau are the two main neuropathological hallmarks of AD [4-6]. The advent of minimally invasive and *in-vivo* imaging of Aβ and tau in the human brain as well as high resolution structural imaging enables investigation of the link between well-documented AD neuropathology and neurodegeneration [7-9]. There are inconsistent reports about the relationship between neurodegeneration and these two pathologies, especially Aβ deposition. Some studies report that an increase in Aβ deposition is associated with neurodegeneration (cortical thinning and/or lower volume) whereas^[10-18]^ other studies report the opposite: higher Aβ deposition is associated with cortical thickening and/or increased volume [19-22]. Several other studies report no relationship between Aβ and neurodegeneration [23-28]. In contrast, most studies consistently report that an increase in tau deposition is associated with a decrease in cortical thickness [12, 20, 21, 25, 27]. More recently, one study reported that the relationship between Aβ pathology and cortical thickness was non-linear and influenced by tau deposition; whereas tau deposition was consistently associated with lower cortical thickness, regardless of Aβ deposition [20]. This study indicated that in participants with normal levels of tau deposition, higher Aβ deposition was associated with an increase in cortical thickness, whereas in participants with higher levels of tau, higher Aβ deposition was associated with a decrease in cortical thickness.

These findings suggest a complex interplay between Aβ, tau, and neurodegeneration which may differ at the early and late stages of AD ^19-21^; thus, measurement of both pathologies might be required for investigating the relationship between AD pathologies and neurodegeneration. Most existing studies consider the relationship between only one of these pathologies and neurodegeneration, which may contribute to the inconsistent findings. Existing studies are also flawed in their cut-point for pathologically negative/positive participants and often define it by clustering the binomial distribution of the global Aβ/tau uptake of the elderly cohort. This approach has resulted in different cut-points across studies and completely disregards the earliest levels of pathophysiological accumulation detected in the majority of participants[29]. In addition, measures of Aβ and tau deposition below the cut-points of established pathologies carry critical information on the earliest regional evolution of these two pathologies and have the potential to track the brain changes due to the earliest pathophysiological consequences.

In this study, we examined 529 participants who had undergone both Aβ and tau ^18^F-MK6240 and ^18^F-Florbetaben/^18^F-Florbetapir positron emission tomography (PET), and magnetic resonance imaging (MRI) scanning, making it possible to study the distinct and joint effects of the Aβ and/or tau pathologies on neurodegeneration (cortical thickness). We derived cut-points for Aβ and tau abnormality by referencing a group of young participants (20 ∼ 40 years) as our healthy controls (HC). Using these redefined cut-points for Aβ and tau deposition based on young subjects who are expected to have no pathological Aβ/tau, we were able to characterize the normal and abnormal levels of Aβ and tau deposition in the older participants. We categorized the older participants into four separate groups: normal Aβ/normal tau (nAβ/nTau), abnormal Aβ/normal tau (aAβ/nTau), normal Aβ/abnormal tau (nAβ/aTau), abnormal Aβ/abnormal tau (aAβ/aTau). We utilized vertex-wise and region-wise multiple linear regression analyses to find the association between cortical thickness and Aβ/tau deposition in each of the four categories. Our primary aim in this study was to assess the distinct associations between cortical thickness and normal/abnormal levels of deposition of each pathology (Aβ and tau) while controlling for the effect of the other pathology (e.g., investigate association of cortical thickness with Aβ deposition, while controlling for tau). Furthermore, we aimed to assess the joint effect of Aβ and tau deposition on cortical thickness in later stages of accumulation and hypothesized that synergy between Aβ and tau leads to unique toxicity that is associated with greater neurodegeneration beyond the individual effects of Aβ or tau deposition alone.

## Methods

### Participants

Data in this study were collected from five separate research cohorts at Weill Cornell Medicine and Columbia University Irving Medical Center. We identified 394 HC and 52 MCI participants 55 years and older (282 females) who underwent three different imaging studies (T1-weighted structural MRI, tau PET, and Aβ PET) within 12 months. All participants consented to participate in their respective studies, and all recruitment/enrollment procedures and imaging protocols were approved by the local institutional review boards. The HC eligible participants underwent standardized medical and neuropsychological evaluations to ensure no neurological or psychiatric conditions, cognitive impairment, major medical diseases, or contraindications based on MRI. The patients with MCI had mini-mental state examination (MMSE) scores of 18–28, a clinical dementia rating (CDR) of 0.5 or 1.0, and the presence of a biomarker associated with AD (either by an Aβ PET scan or cerebrospinal fluid (CSF) analysis showing a positive Aβ42, tau, and/or phospho-tau protein181).

### Image acquisition protocols

All magnetization-prepared rapid gradient-echo (MP-RAGE) scans were acquired with 3.0 Tesla MRI scanners. Each participant first underwent a scout localizer to determine the position and set the field of view and orientation, followed by high resolution MP-RAGE image with TR /TE = 2300-3000/2.96-6.5 ms, flip angle = 8-9°; field of view = 25.4-26 cm, matrix size = 256×256, and 165-208 slices with 1 mm thickness.

Tau PET imaging for all the participants was done using ^18^F-MK6240. Each participant’s vital signs were recorded before and after tracer-injection. An intravenous catheter (IV-line) was inserted into the arm, and an injection of 185 MBq (5 mCi) ± 20% (maximum volume 10 mL) was administered as a slow single IV bolus at 60 seconds or less (6 secs/mL max). Post-injection saline flush of the IV line was not allowed. A low-dose computed tomography (CT) scan for the attenuation correction of the PET data was acquired. Starting at 80-120 minutes’ post-injection, brain images were acquired in 6 × 5-minute frames over a period of 30 minutes. If considered inadequate, the participant underwent an additional 20 minutes of continuous imaging.

For Aβ imaging, HC and MCI participants underwent ^18^F-Florbetaben and ^18^F-Florbetapir PET scans, respectively. Each participant’s preparation for the scans consisted of an IV catheterization, followed by the injection of 8.1mCi ± 20% (300 MBq) of the tracer administered as a slow single IV bolus at 60 seconds or less (6 secs/mL max). There were two separate post injection imaging start times for the acquired Aβ-specific scans. Participants were scanned 45-90 minutes after tracer injection. A low-dose CT scan for attenuation correction of the PET data was also acquired. Brain images for each of these PET scans were acquired in 4 × 5-minute frames over a period of 20 minutes.

### Quantification of structural imaging data

The T1-weighted structural scans were reconstructed using FreeSurfer (http://surfer.nmr.mgh.harvard.edu) automated segmentation and cortical parcellation software package [30, 31]. FreeSurfer segments the cortex into 33 different gyri/sulci-based regions in each hemisphere according to the Desikan-Killiany atlas [32], and subcortical segmentation and calculates the cortical thickness at each vertex at millimeter-by-millimeter resolution. The vertex-wise data are not constrained to the pre-defined regions of interest (ROI)s and can be transferred to standard space using surface-based non-linear registration. We utilized vertex-wise data to detect effects that are smaller in size over two or more pre-defined ROIs. The transfer of all neuroimaging data to standard space (MNI152) was also required for group analyses. We used advanced normalization tools (ANTs) [33] to transfer the participant’s native space voxel intensities to the MNI space for any voxel-wise group comparison. For vertex-wise analysis, we projected the surface base reconstructed PET to the surface of MNI152 using the spherical surface registration in FreeSurfer.

### Quantification of molecular imaging data

To process the Aβ and tau PET scans, a fully automatic in-house developed quantification method was used. This method has already been used in numerous studies and validated using histopathological data [34-38]. Briefly, dynamic PET frames (six frames in tau PET and four frames in Aβ PET) are first aligned to the first frame using rigid-body registration and averaged to generate a static PET image. The PET image was then registered with the CT image and merged to obtain a composite image in the PET static space. Next, the structural T1 image in FreeSurfer space was also registered to the same participant’s CT/PET composite image using normalized mutual information and six degrees of freedom to obtain a rigid-body transformation matrix. The FreeSurfer regional masks in static PET space were used to extract the regional PET data. The standardized uptake value (SUV), defined as the decay-corrected brain radioactivity concentration normalized for injected dose and body weight, was calculated; it was normalized to cerebellum gray matter to derive the standardized uptake value ratio (SUVR). The FreeSurfer cortical and subcortical regions, as well as vertex-wise surface reconstruction, were used in the native space analysis of the PET data [39]. Finally, all vertex-wise quantification of Aβ, tau, and surface-based probabilistic atlases are generated by FreeSurfer’s reconstructed surfaces. In addition, we quantified the average of each region’s SUVR for region-based analyses.

The Aβ and tau uptakes in AD-relevant regions were also quantified. For Aβ, the global SUVR was calculated by targeting regions of interest including frontal, parietal, temporal, anterior and posterior cingulate, and precuneus regions. Tau target ROIs were in temporal lobe AD-related regions including the fusiform, amygdala, parahippocampal gyrus, entorhinal, inferior temporal, and middle temporal regions. To capture the early effect of categorizing the participants based on tau PET, we defined the medial temporal lobe (MTL) tau with ROIs, including the entorhinal and parahippocampal gyrus Tau deposition has been shown to accumulate first in these two regions [40, 41]. Global Aβ and global tau, and MTL tau SUVRs, were utilized to find the cut points for Aβ and tau deposition abnormality.

### Partial volume correction

We developed a simple but effective anatomy-driven partial volume correction (PVC) technique with a sufficiently powered normative reference group (age: 20 ∼ 40). Each gray matter voxel’s uptake is a combination of actual binding in that location and the spill-in from white-matter/meningeal non-specific binding for Florbetaben/MK6240 scans. Using Florbetaben/MK6240 scans from 62/21 healthy young (years<40) participants’ Aβ/tau images, we estimated the white matter/meningeal spill-in signal. Since the majority of young (years<40) and healthy participants are not expected to have any Aβ/tau accumulation, any gray matter uptake in these participants can be considered a result of spill-in from the non-specific binding of adjacent regions. We used the white matter/meninges mask for each young participant to extract the spatial distribution of the non-specific binding within these regions. Convolving this mask with the scanner point-spread function simulates the effect of a spill-in signal perfectly since within the gray matter regions the correlation between actual uptake and the synthesized spill-in was more than 80%. Then, we fitted a linear regression model for each voxel to predict the synthesized spill-in’s gray-matter. We extracted the white matter/meninges uptake in the test participants and convolved it with a scanner point spread function to get the synthesized spill-in for the gray-matter voxels. Using the fitted model parameters at each voxel, spill-in amount (synthesized spill-in) was estimated. Finally, we subtracted the actual uptake with the synthesized amount to completely remove the artifacts.

### Subject categorization

In this study, we categorized the older participants by determining cut-points based on young participants’ global Aβ and global/MTL tau uptakes. We first obtained the young participants’ global Aβ and global/MTL tau distributions, which were normally distributed according to the Shapiro-walk test: P>0.22. Then, by using the 95^th^ percentile of the fitted normal distribution, we calculated the cut-points for abnormal global Aβ and global/local tau uptakes. Note that MTL tau uptake was to detect the early stages of abnormality. Therefore, the abnormality of the tau was determined when the global or MTL tau uptake level was higher than their associated cut-points. Using this categorization technique, each participant can be categorized as abnormal (a) and normal (n), resulting in four groups: 1-nAβ/nTau: participants with neither abnormal global Aβ nor global/MTL tau pathologies. 2 - aAβ/nTau: participants who have normal global/MTL tau but abnormal Aβ; 3-nAβ/aTau: participants who have abnormal global or MTL tau but normal Aβ pathologies; 4-aAβ/aTau: participants who have abnormal Aβ and global or MTL tau pathologies.

### Statistical analysis

After categorizing the older participants with the defined cut-points based on healthy young participants, the probabilistic atlas was obtained for visualizing the pattern of Aβ and tau deposition on the surface of the brain in each category. We thresholded and then binarized each vertex of Aβ and tau uptakes by the obtained cut-points of 1.256 and 1.150, respectively (higher than cut-point= 1, and lower than cut-point= 0). Finally, we computed the probability of observing the abnormal Aβ/tau (based on cut-points) across participants of each category. In addition, inter-regional correlations between Aβ and tau uptakes across the different categories were calculated.

To assess the association between global Aβ/tau and cortical thickness in the normal and abnormal levels of deposition, the first multiple regression model was applied to each brain vertex while age, gender, and ICV were controlled as covariates: *Vertex-Wise Cortical Thickness ∼ β*_*0*_ *+ β*_*1*_ *Global Aβ + β*_*2*_ *Global Tau + β*_*3*_ *Age + β*_*4*_ *Gender + β*_*5*_ *ICV + e*. Then, we generated the Aβ/tau statistical maps (t-test) to visualize vertices with significant associations between cortical thickness and Aβ/tau deposition. The second analysis was applied to test the regional association between Aβ/tau deposition and cortical thickness in 68 target regions. To exclude the effects of age, gender, and ICV, we first residual them from the regional cortical thickness. It is noteworthy that in each pathology’s relationship (Aβ or tau), the other one also residual out to find the pure effect of the targeted pathology. For example, to find the association between regional thickness and Aβ deposition, the regional tau is considered the first residual: *Regional Cortical Thickness ∼ β*_*0*_ *+ β*_*1*_ *Regional Aβ or Regional Tau + β*_*3*_ *Age + β*_*4*_ *Gender + β*_*5*_ *ICV + e*. Finally, we ran a second regression model to find the relationship between the target regions’ cortical thickness and each pathology’s regional uptake (*Residuals Regional Cortical Thickness ∼ β*_*0*_ *+ β*_*1*_ *Regional Aβ or Regional Tau + e*).

The joint effect of Aβ and tau deposition was explored by a resonance measure, in which resonance is the spatial correlation between the amount of Aβ and tau uptake across gray matter vertices. The resonance measure shows the synergetic relationship between Aβ and tau uptake in the cerebral cortex by calculating the vertex wise correlation. Each participant has one resonance measurement, which shows how much the spatial pattern of Aβ uptake in the cerebral cortex is associated with the spatial pattern of tau uptake. The multiple linear regression model was then performed on each vertex to determine the joint effect of Aβ/tau deposition (resonance) on cortical thickness while controlling for the effects of age, gender, ICV, and global Aβ/tau: *Cortical Thickness ∼ β*_*0*_ *+ β*_*1*_ *Global Aβ + β*_*2*_ *Global Tau + β*_*3*_ *Resonance + β*_*4*_ *Age + β*_*5*_ *Gender + β*_*6*_ *ICV + e*. We also performed regional regression to see whether the resonance by itself was associated with regional cortical thickness in the four categories.

All statistical analyses and their visualization in this study were performed using Python. The main numeric modules that were utilized were NumPy and Matplotlib [42, 43]. The Student’s t-tests, Chi-square, and ANOVA tests were performed using the SciPy statistical package (v6.1.1) [44]. The vertex-wise false discovery rate (FDR) with p-value=0.05 correction was performed for vertex-wise analyses in FreeSurfer Qdec[45]. For family-wise error correction of regional associations, we performed a permutation test. We randomly shuffled the independent variable 10,000 times to find an empirical null distribution for the t-value of regression analysis. Finally, the family-wise error rate-corrected t-value was calculated based on the 5^th^ and 95^th^ percentiles of the fitted normal distribution for negative and positive t-values.

## Results

### Subjects’ categorization

In this study, for the first time, we categorized older participants in reference to the global SUVR of the Aβ/tau signal observed in a young population. Fig. 1 depicts the distribution of the young (in orange) and older (in blue) participants’ global Aβ (Fig. 1a), global tau (Fig. 1b), and MTL tau (Fig. 1c) uptakes. As depicted, the 95^th^ percentile of the normal distribution fitted to the young participants (the black dotted line) is used as the cut-points for distinguishing abnormal global Aβ (Fig. 1a) and global/MTL tau (Figs. 1b/1c). The global Aβ standardized uptake value ratio (SUVR) cut-point was 1.256 and the global and local tau SUVR cut-point were 1.150 and 1.110, respectively.

**Fig. 1:**
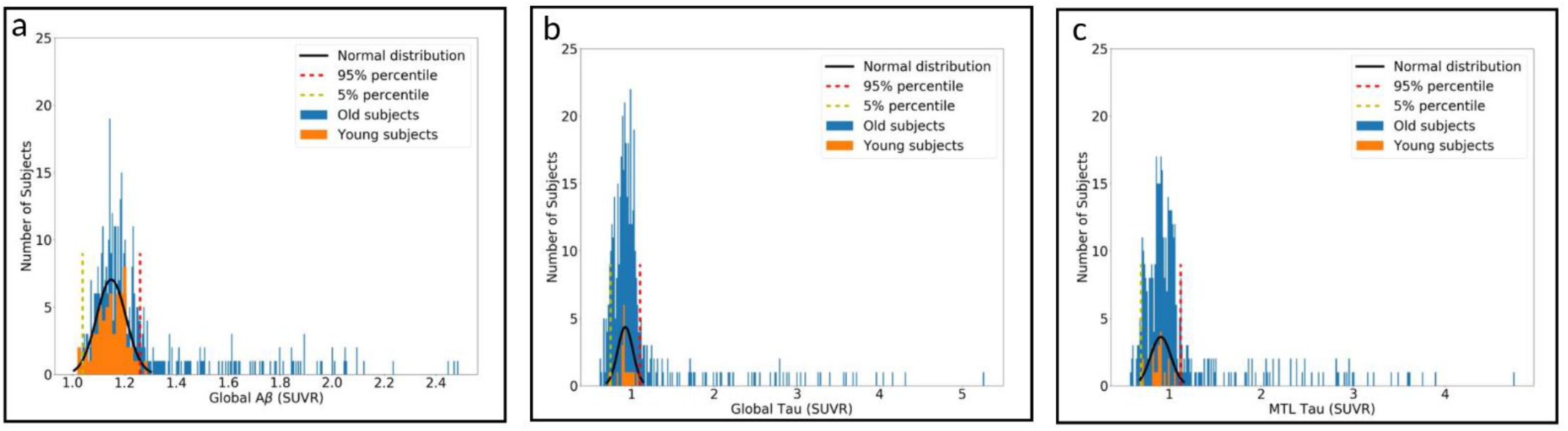
Distribution of younger (in orange) and older (in blue) participants’, (a) global Aβ, (b) global tau, and (c) MTL tau. The fitted normal distribution (in black), 95^th^ percentile (red dotted line), and 5^th^ percentile (yellow dotted line) overlaid on the distributions of the participants.

Table 1 illustrates the number of participants and their demographic in each of the four categories in this study. Our categorization results indicate that ∼52% of participants fall in the nAβ/nTau group, covering 58% of HC participants and 5.8% of MCI patients. While almost 75% of the participants with mild cognitive impairment (MCI) fall in the aAβ/aTau group the remaining 25% of MCI participants are distributed between the other three groups (5.8% in nAβ/nTau, 5.8% in nAβ/aTau, and 13.4% in aAβ/nTau).

**Table 1.** Cohort demographics

| | Young Tau<br>PET Participa<br>nt<br>(N=21) | Young Aβ<br>PET Particip<br>ant<br>(N=62) | Old Aβ/Tau<br>PET<br>participant<br>(n=446) | nAβ/nTau<br>(n=232) | aAβ/nTau<br>(n=46) | nAβ/aTau<br>(n=96) | aAβ/aTau<br>(n=72) | Four<br>group difference<br>(P-value) |
| --- | --- | --- | --- | --- | --- | --- | --- | --- |
| **Age (years)** | 29.40<br>(5.79) | 27.40<br>(5.10) | 66.25<br>(5.29) | 64.79<br>(3.55) | 66.84<br>(5.17) | 67.20<br>(5.90) | 69.69<br>(7.11) | P<0.0001 |
| **Sex (M/F)** | 8/13 | 24/38 | 164/282 | 94/138 | 16/30 | 32/64 | 25/47 | 0.573 |
| **HC/MCI** | 21/0 | 62/0 | 394/52 | 229/3 | 39/7 | 93/3 | 33/39 | P<0.0001 |
| **Global Aβ SUVR** | - | 1.14<br>(0.05) | 1.25<br>(0.24) | 1.14<br>(0.07) | 1.37<br>(0.13) | 1.15<br>(0.06) | 1.71<br>(0.29) | P<0.0001 |
| **Global Tau SUVR** | 0.92<br>(0.09) | - | 1.20<br>(0.66) | 0.94<br>(0.09) | 0.95<br>(0.09) | 1.20<br>(0.34) | 2.23<br>(1.11) | P<0.0001 |

Abbreviations: HC: healthy control, MCI: mild cognitive impairment, M: male, F: female, PET: positron emission tomography, nAβ /nTau: normal Aβ /normal tau, aAβ /nTau: abnormal Aβ /normal tau, nAβ /aTau: normal Aβ /abnormal tau, aAβ /aTau: abnormal Aβ /abnormal tau, SUVR: standardized uptake value ratio

Fig. 2a illustrates the probability of observing the Aβ and tau pathologies (SUVR > global cut-point) at each vertex throughout the entire cerebral cortex in the four categories of participants given in Table 1. The probabilities are overlaid on a semi-inflated cortical surface of the MNI152 template and color-coded with a heat color map where the darker red and red indicate lower probabilities, and the bright red and yellow indicate higher probabilities. These results emphasize the existence of an abnormal level of local/regional uptake of Aβ and/or tau in participants with normal levels of global Aβ and/or tau. As such, even participants in the first category (nAβ/nTau) possess some abnormal level of local Aβ and/or tau deposition in some specific regions. Also, participants in the last category (aAβ/aTau) have abnormal Aβ and tau deposition extensively throughout the brain.

**Fig. 2:**
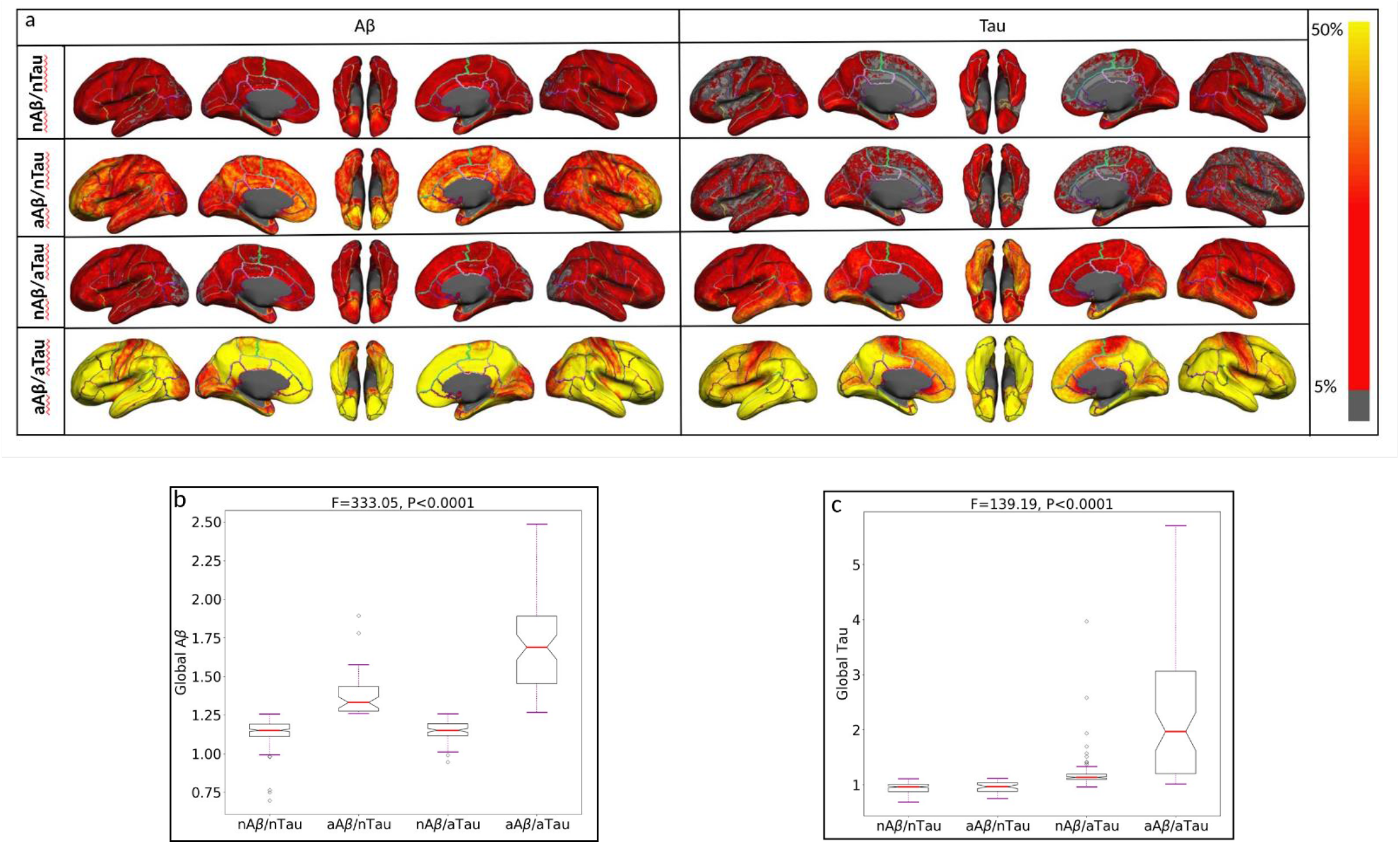
(a) Illustrating the vertex-wise probabilistic atlas of Aβ (left column) and tau (right column) pathologies throughout the entire cerebral cortex obtained in four participant categories. (First row, nAβ/nTau; second row, aAβ/nTau; third row, nAβ/aTau; and fourth row, aAβ/aTau). The probability of observing Aβ and tau at each vertex is color-coded with a heat color map and overlaid on a semi-inflated cortical surface of the MNI152 template. **(b)** Boxplots compare the distribution of global Aβ in four categories of participants. **(c)** Boxplots compare the distribution of global tau in four participant categories.

The distribution of the global Aβ and global tau uptakes in the four groups are depicted by boxplot in Fig 2b and 2c, respectively. As expected, a one-way ANOVA comparing the four participant groups found a significant difference between global Aβ (F=333.05, P <0.0001) and global tau (F=139.19, P <0.0001). The post-hoc pair-wise Student’s t-test also shows a substantial difference in the global Aβ deposition (t>7.19, p<0.0001) between all paired categories except between the two normal Aβ groups (nAβ/nTau and nAβ/aTau). Furthermore, the Student’s t-test shows a significant difference between all pairs of categories (t>4.79, p<0.0001) in global tau except between the two normal tau groups (nAβ/nTau and nAβ/aTau).

The inter-regional Aβ and tau association were computed separately for each of the four distinct categories of subjects and depicted in Fig. 3a-d using a color-coded cross-correlogram. Each element of the cross-correlogram is color-coded with a heat map and represents a subject-wise correlation between the x-axis region’s Aβ uptake and the y-axis region’s tau uptake. The red color indicates a correlation value equal to 1, and the blue color indicates a correlation value equal to −1. Fig. 3a-c illustrate the early stage of association between Aβ and tau accumulation in nAβ/nTau, aAβ/nTau, and nAβ/aTau groups respectively. It is apparent that there are weak but important correlations in some regions (yellow color) that might show an early association between Aβ and tau. On the other hand, in Fig. 3d, the correlation gets strongly positive in almost all brain regions. These results demonstrate the hitting point of the joint effect (resonance effect between Aβ and Tau pathologies) that happens in the late stage of accumulation which strongly accelerates the synergic effect of these two pathologies. In other words, these results suggest that while Aβ and tau initially start accumulating in different regions of the cortex independently, their accumulation spreads to the entire cortex in the aAβ/aTau group highlighting the possibility of synergy between them, henceforth referred to as *resonance*.

**Fig. 3:**
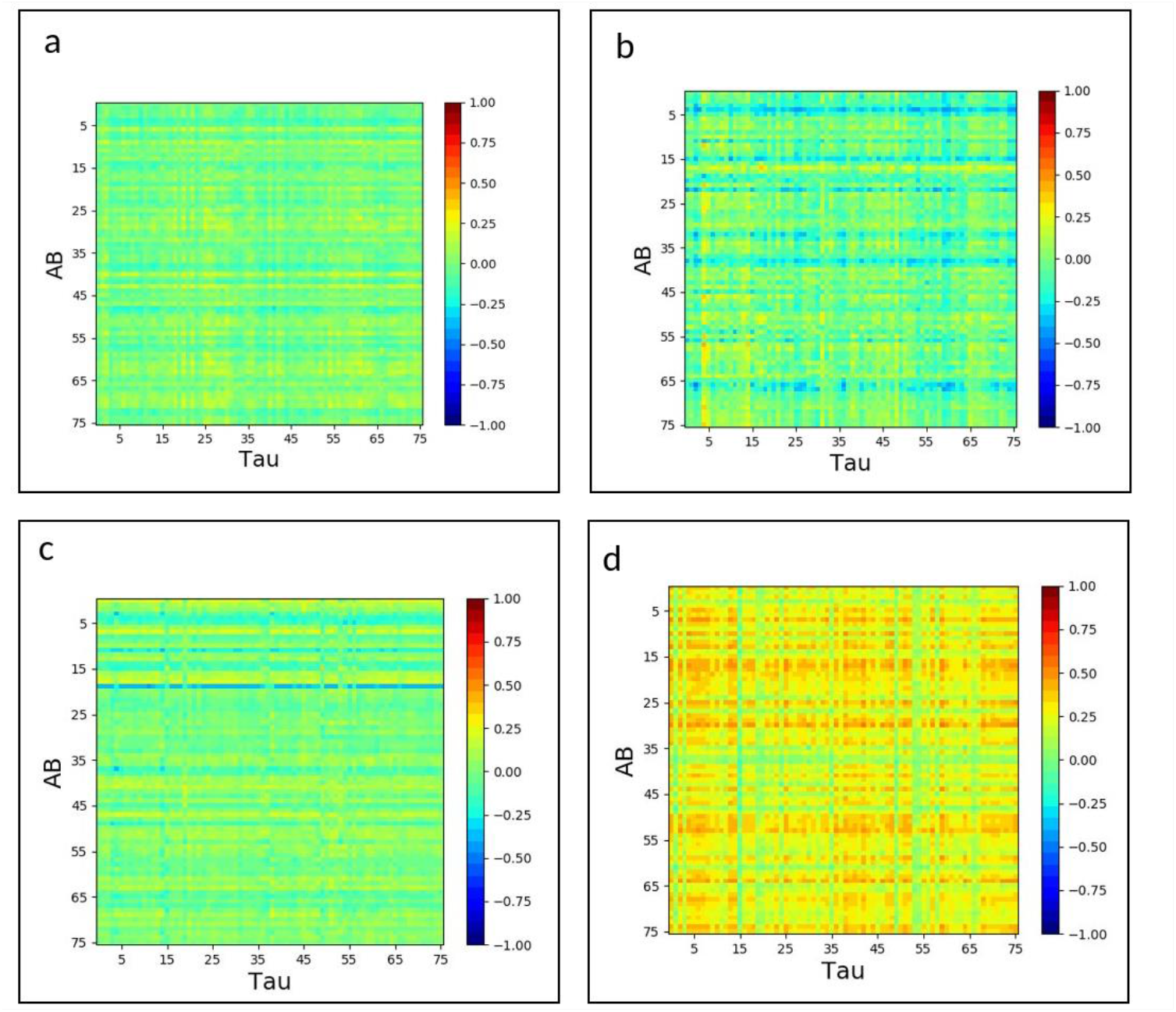
Inter-regional cross-correlogram between Aβ and Tau accumulations for (a) nAβ/nTau, (b) aAβ/nTau; (c) nAβ/aTau, and (d) aAβ/aTau groups in 76 cortical regions. The correlation is color-coded with a heatmap, and the red color indicates a correlation value equal to 1, and the blue color indicates a correlation equal to −1

### AD pathologies and vertex-wise cortical thickness

Using a vertex-wise multiple linear regression model, we explored the effect of Aβ and tau deposition on cortical brain atrophy as a measurement of neurodegeneration. Fig. 4 demonstrates the results of this analysis, performed separately for each category of participants after multiple comparisons correction (the uncorrected results show in Fig. S1). It depicts all vertices with a significant association between global Aβ (left column)/tau (right column) deposition and cortical thickness when controlling for the other pathology (tau/Aβ), age, gender, and intracranial volume (ICV). As shown, an increase in global Aβ deposition is associated with an increase in cortical thickness at the normal level of global deposition (nAβ/nTau and nAβ/aTau) regardless of tau level. It is noteworthy that the regions showing increased cortical thickness in association with a normal level of global Aβ deposition in the medial temporal lobe are the same regions where the earliest tau deposition has been reported [40]. In contrast to Aβ, tau deposition shows a differential trend depending on the level of deposition. As shown in Fig. 4, at normal levels of global or MTL tau deposition, an increase in global tau accumulation is associated with an increase in cortical thickness. By contrast, at abnormal levels of global tau deposition, an increase in global tau accumulation is strongly associated with a decrease in cortical thickness, regardless of Aβ level.

**Fig. 4:**
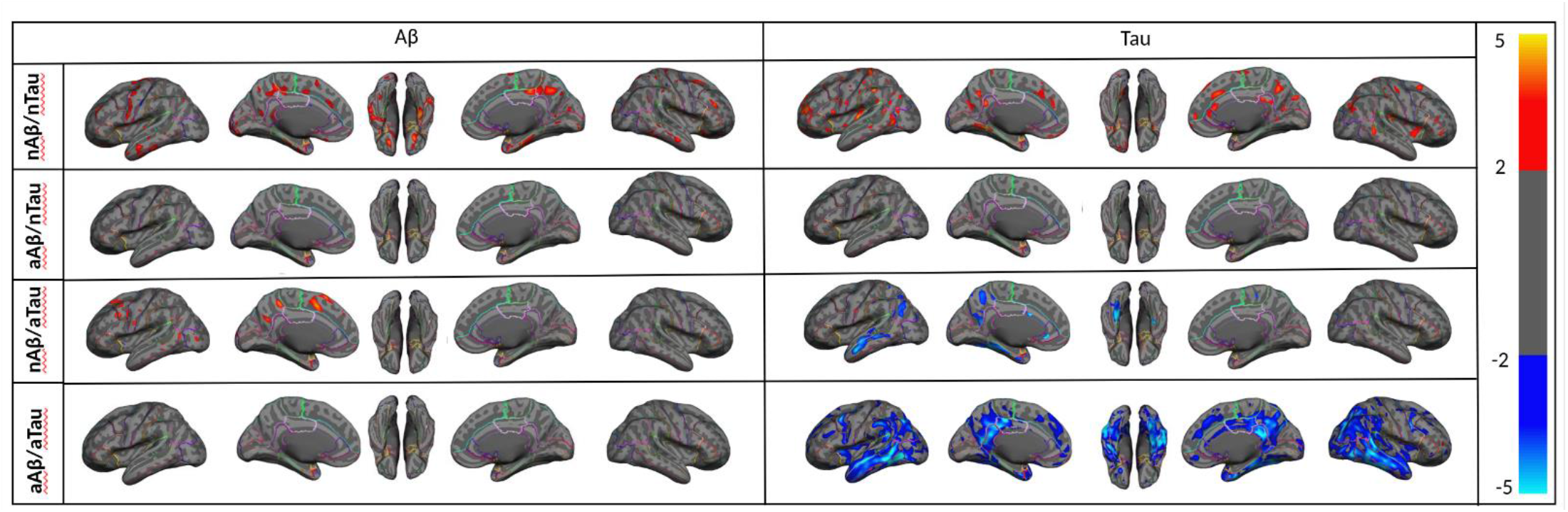
Vertex-wise statistical map (t-value) of association between global Aβ (left column), global tau (right column) pathologies, and cortical thickness throughout the entire cerebral cortex obtained in four categories of participants. (First row, nAβ/nTau; second row, aAβ/nTau; third row, nAβ/aTau; and fourth row, aAβ/aTau). The t-value at each vertex is color-coded with red to yellow colors representing increasing positive t-values and blue to light blue representing decreasing negative t-values and overlaid on the semi-inflated cortical surface of the MNI152 template. The association between global Aβ, global tau pathologies, and cortical thickness survived after multiple comparison corrections with FDR.

### Regional AD pathologies and cortical thickness

Fig. 5a depicts associations between regional cortical thickness and mean Aβ/tau deposition in 68 regions while controlling for the other pathology (tau/Aβ), age, gender, and ICV. Fig. 5b illustrates scatter plots overlaid with the association between the residual average cortical thickness and Aβ deposition in the left entorhinal and right posterior cingulate as two target regions for Aβ. Fig. 5c shows residual cortical thickness and tau deposition in the left entorhinal and left insula as two target regions for tau. The results shown in Fig. 5a-c are corrected for multiple comparisons correction (FWE); uncorrected results are shown in Fig. S2. As expected from previous results, at normal Aβ levels, an increase in regional Aβ uptake is significantly associated with an increase in regional cortical thickness in the entorhinal cortex for nAβ/nTau (t > 3.241) and posterior cingulate cortex for nAβ/nTau, aAβ/aTau, and nAβ/aTau (t > 3.889, 3.028, and 3.805, respectively; Fig. 5b). Interestingly, there are several subjects in the nAβ/nTau group with abnormal regional Aβ uptakes >1.25 (Fig. 5b). These participants might drive the aforementioned perceived increase in regional cortical thickness. Of note, this relationship masked out when the regression analysis was performed with global Aβ (Fig. 4). At normal levels of global tau, an increase in regional tau uptake is associated with a significant increase in regional cortical thickness in the insula (t> 3.202; Fig. 5c). By contrast, at abnormal levels of global tau, an increase in regional tau uptake is associated with a significant decrease in cortical thickness in the entorhinal cortex for nAβ/aTau and aAβ/aTau (t-value< −4.372, −2.80, respectively), and the insula (t-value < −4.284; Fig. 4c). These results emphasize that even at normal levels of global Aβ deposition (nAβ/nTau and nAβ/aTau) there are surprisingly regions with high levels of regional Aβ deposition, like the entorhinal cortex, that are associated with an increase in cortical thickness. By contrast, regardless of Aβ levels, regional abnormal tau levels are associated with a strong reduction in cortical thickness in several brain regions.

**Fig. 5:**
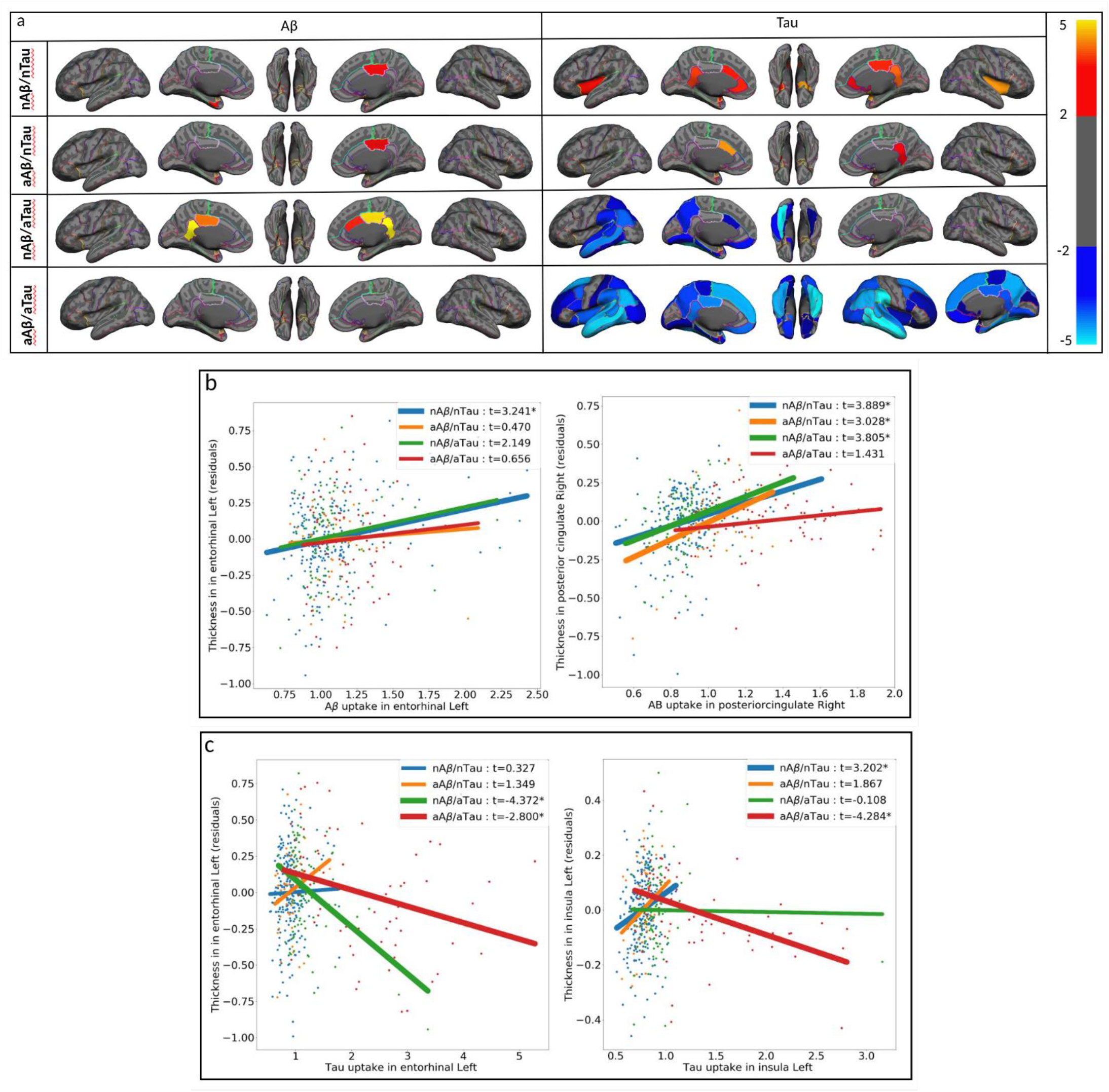
(a) Region-wise statistical map (t-value) of association between regional Aβ (left column), regional tau (right column) pathologies, and regional cortical thickness throughout 68 ROIs obtained in four categories of participants (First row, nAβ/nTau; second row, aAβ/nTau; third row, nAβ/aTau; and fourth row, aAβ/aTau). The t-value at each region is color-coded with red to yellow colors representing increasing positive t-values and blue to light blue representing decreasing negative t-values and overlaid on the semi-inflated cortical surface of the MNI152 template. (b) The regional multiple regression analysis results in the association between residual regional cortical thickness and Aβ in two target regions: left entorhinal and right posterior cingulate. (c) The regional multiple regression analysis results of the association between residual regional cortical thickness and tau, in two target regions: left entorhinal, and left insula. Association between regional Aβ, tau pathologies and cortical thickness survived after family-wise error correction. In Fig. 4b-c the survived associations relationships are shown with thicker lines and *.

### AD pathologies and cortical thickness in young participants

While some studies report that an increase in Aβ deposition is associated with an increase in cortical thickness, such a relationship has never been reported for tau and neurodegeneration. Since the positive relationship between early Aβ/tau deposition and cortical thickness is obtained based on global Aβ and global tau uptake distributions from young participants, it is reasonable to assess whether such a relationship also holds for the young groups. We performed the same analysis to assess the relationship between vertex-wise cortical thickness and global Aβ and tau uptakes in 62 and 21 young participants, respectively. It is noteworthy that we did not have access to both Aβ and tau imaging for all young subjects; so, for the analysis of each target pathology (Aβ or tau), we did not control the other one as a separate covariate. Fig. S3 shows the corrected association between cortical thickness and global Aβ in 62 young participants. As shown in Fig. S3, no association survived after FDR correction. In Fig. S4 we performed the same analysis for global tau using 21 young subjects. The corrected vertex-wise results in Fig. S4 illustrate that no association survived after FDR correction. The lack of any association between global Aβ/global tau deposition and cortical thickness in the young participants can be used as solid evidence that our results in the old participants are reporting a true effect of early accumulations on cortical thickness. Also, reject any hypothesis about the effects of the blood perfusion (as a confounder for PET images) on the results of old participants.

### The joint effect of AD pathologies on Neurodegeneration

Once the unique and distinct effects of each AD pathology on cortical thickness were demonstrated, we aimed to assess any remaining joint effect of Aβ and tau deposition on the cortical thickness that is not accounted for by their linear effects. We have shown previously [46] and in this paper that while Aβ and tau are initiated independently at different brain regions, they later progress to a state of “resonance”. Therefore, we also aimed to assess whether the resonance between Aβ and tau is also associated with neurodegeneration beyond what we have already accounted for in the linear effects of Aβ and tau. Fig. 6 shows the association between cortical thickness and resonance in the aAβ/aTau group after FDR correction, controlling for age, gender, ICV, global Aβ, and global tau. The associations between cortical thickness and resonance in the three other groups (nAβ/nTau, aAβ/nTau, and nAβ/aTau) did not survive after FDR corrections (Fig. S5 shows the uncorrected associations in each of the four different groups). As shown in Fig. 6, increased resonance at higher levels of deposition is strongly associated with a decrease in cortical thickness. This joint effect (resonance) is beyond the Aβ and tau linear additive effects since we controlled for global Aβ and tau uptakes in the multiple regression analyses. Fig. 7a depicts the joint effect of regional Aβ and tau on regional cortical thickness in the aAβ/aTau group. Whereas the other three groups’ regional associations did not survive after we applied family-wise error correction (permutation test). Fig. S6 shows the uncorrected associations in each of the four different groups. Increased resonance at abnormal levels of deposition (aAβ/aTau group) is strongly associated with a decrease in cortical thickness across several regions in the brain like the parahippocampal left (t<-2.359) and caudal anterior cingulate right with (t<-2.661; Fig. 6b). Altogether, these results highlight the non-additive feature of the joint effect of Aβ and tau on cortical atrophy at abnormal levels of deposition (aAβ/aTau group). These results also highlight that the resonance between these two pathologies may accelerate the effect of the pathologies on cortical thickness beyond the additive effects of Aβ and tau.

**Fig. 6:**
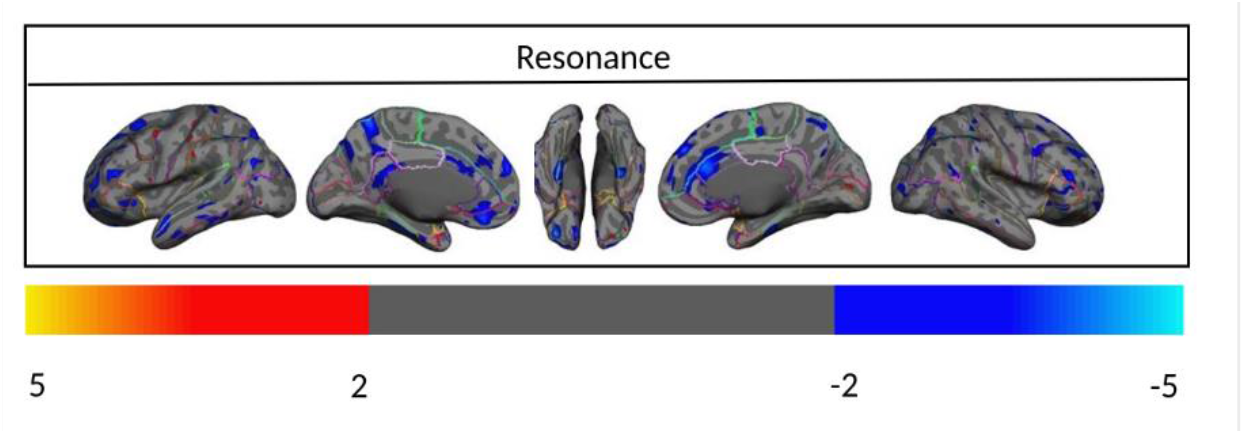
(a) Vertex-wise statistical map (t-value) of the association between resonance and cortical thickness throughout the entire cerebral cortex obtained in four categories of participants in aAβ/aTau group. The t-value at each vertex is color-coded with a heatmap where red to yellow shading represents increasing positive t-values and blue to light-blue shading represents decreasing negative t-values overlaid on the semi-inflated cortical surface of the MNI152 template. Association between resonance and cortical thickness survived after multiple comparison corrections with FDR.

**Fig. 7:**
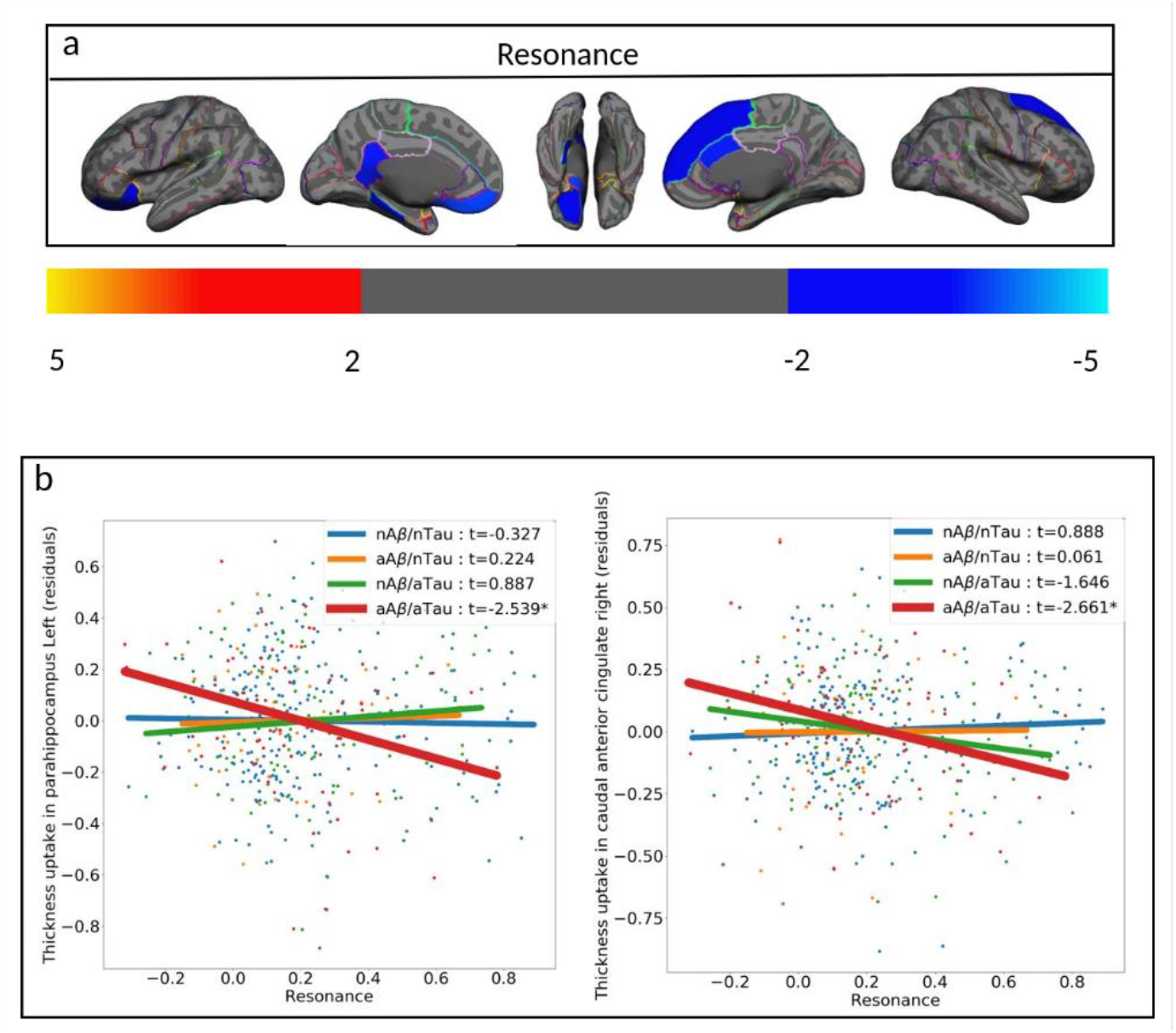
(a) Region-wise statistical map (t-value) of the association between resonance and cortical thickness throughout 68 ROIs obtained in aAβ/aTau group. The t-value at each region is color-coded with a heatmap where red to yellow colors represent increasing positive t-values and blue to light-blue represents decreasing negative t-values overlaid on the semi-inflated cortical surface of the MNI152 template. (b) The regional multiple regression analysis results demonstrate the association between regional cortical thickness and resonance in two target regions: left parahippocampal, and right caudal anterior cingulate. Association between resonance and cortical thickness survived after family-wise error correction. In Fig. 5b the survived associations relationships are shown with thicker lines and *.

## Discussion

In this study, we introduced a new categorization method for older participants based on the distribution of global Aβ and tau deposition in young (<40 years old) participants. This approach offers important insights into the earliest stage of AD pathologies and their spatial patterns of accumulation and can be used to study both the distinct and joint associations between Aβ/tau deposition and neurodegeneration. In this study, we propose that the effects of Aβ and tau pathologies in the brain can be divided into two detached subtypes: 1) the distinct effects of Aβ and tau, and 2) the joint effects of Aβ and tau. We first demonstrated that normal levels of global Aβ deposition were associated with an increase in cortical thickness when we controlled for tau deposition and other confounders (age, gender, and ICV). On the other hand, when global Aβ deposition and other covariates were controlled, normal levels of global tau deposition were associated with an increase in cortical thickness, while abnormal levels of global tau deposition were associated with a decrease in cortical thickness. Next, the regional association between these two pathologies and cortical thickness provided supporting evidence of the association between normal levels of Aβ deposition and an increase in thickness, particularly in the entorhinal cortex, where tau deposition has been shown to initiate [47, 48]. In addition, even small amounts of abnormal tau deposition were associated with decreased cortical thickness. To provide further evidence, we performed the same analyses on young participants and found no significant association between normal levels of Aβ or tau and cortical thickness in young participants. Finally, we investigated the joint effect of Aβ and tau deposition on cortical thickness, beyond their linear additive effects on the brain at abnormal pathology levels.

Several studies have investigated the associations between Aβ and tau pathologies and neurodegeneration. However, there is little consensus about the association between these two pathologies and neurodegeneration in the brain, as studies have variously reported an Aβ-related decrease, increase, and even no change in cortical thickness [10-18, 20, 21, 25, 27]. Previous studies used several different quantitative approaches to find global SUVR cutpoints for Aβ and tau abnormality. These were typically higher than 1.3 and based on different PET tracers [49-53]. Existing studies mostly relied on categorizing participants into negative and positive groups using cut-points of Aβ and tau uptakes that were derived solely based on the older populations. Alternatively, since HC participants are often used as the best discriminating biomarker of the disease, some studies determined cut-points specifically to discriminate between subjects with established Aβ pathology based on MCI and/or AD patients and HC participants. However, accumulations of Aβ and tau have also been reported in normal aging [54], suggesting that participants who fall below the typical cut-point determined in older cohorts may have significant AD pathology in their brain without exhibiting clinical symptoms [29]. Lowering these cut-points using a younger reference sample increases sensitivity to the early stages of Aβ and tau accumulations, which may be biologically important. To our knowledge, the present study is the first that uses young participants to define the cut-point to categorize older participants, thus allowing us to study the effects of early accumulation of AD pathologies on neurodegeneration even in normal aging brains.

Another factor that might have contributed to the inconsistent findings in the literature is the lack of consideration for the interaction between Aβ and tau deposition. Despite the already reported interaction between Aβ and tau pathologies [55, 56], and much effort devoted to investigating the distinct association of each AD pathology and neurodegeneration [20, 21], most existing studies failed to control the effect of one pathology when investigating the effect of the other pathology on neurodegeneration. In this study, by adding both pathologies as independent variables into the multiple regression analysis, we always controlled for the effects of one pathology when investigating the distinct effects of the other pathology on neurodegeneration.

The results of this study, as well as other studies [19-22], provide evidence for a differential relationship between AD pathologies and cortical brain atrophy. One study showed that higher Aβ deposition was associated with an increase in cortical thickness in HC participants when tau deposition was at a normal level [20]. An increase in cortical thickness was also reported by a recent study in AD patients, which showed that higher Aβ was associated with an increase in cortical thickness in a region that otherwise shows AD-related atrophy [4]. On the other hand, several other studies reported a decrease in cortical thickness associated with tau deposition [12, 20, 21, 25, 27]. These findings, and those of our study, highlight the distinct roles of Aβ and tau in the neurodegenerative process and indicate that the two AD pathologies differentially correlate with cortical atrophy. However, to our knowledge, until this study, no prior research had reported an increase in cortical thickness at normal levels of tau accumulation. As such, the extent to which these pathologies may have an increasing cortical thickness remains unclear and future studies are needed to determine the biological relationship between increasing cortical thickness and AD pathologies. Here we offer three possibilities that might explain how AD pathologies could be associated with an increase in cortical thickness. First, cortical thickening might be driven by brain hyperactivation to compensate for the disruptive effects of pathologies [57-60]. AD pathologies in the brain can also change the balance between synaptic excitation and inhibition [61] leading to cellular hyperactivity which then may cause cortical enlargement. Second, cortical enlargement, particularly due to Aβ deposition, can be a result of inflammation driven by the neuroimmune response that causes local fluid increases (inflammation) [62-64]. Third, the Aβ plaques accumulated within the cortical gray matter also occupy space which can result in an increase in the measurements of cortical thickness and subsequently can induce a false correlation between Aβ deposition and cortical thickness [20]. Last but certainly not least, the thicker cortical regions may require higher blood perfusion which has been shown to be a confounder for PET images, particularly for ^18^F tracers.

Prior studies have suggested the possibility that Aβ and tau deposition start to interact with each other to propagate the pathologies to the entire cerebral cortex^35^. It has also been proposed that abnormal tau and Aβ accumulations are strongly associated with each other in the limbic and neocortical regions [55, 65]. A longitudinal study also reported that, compared to lower amounts of Aβ accumulation, higher amounts of Aβ accumulation drive a greater acceleration of tau accumulation, [66]. The same group also reported that the rate of tau accumulation in HC participants with an abnormal level of Aβ deposition was significantly greater than the rate of tau accumulation in participants with a normal level of Aβ. Using a publicly available database, Alzheimer’s Disease Neuroimaging Initiative (ADNI), we have also demonstrated that spatially overlapping pathologies within the default mode network (DMN) are a robust biomarker for predicting conversion from HC to MCI and MCI to AD [46]. We also suggested that the maladaptive *resonance* between Aβ and tau starts when the two pathologies contaminate the same cortical brain regions and/or networks, and introduced the *spatially-overlapping-insults* [46] hypothesis. According to our hypothesis, when the effects of Aβ and tau overlap and synergize, the resonance between the two pathologies can be detected by assessing the spatial similarity between their pattern of accumulation throughout the entire cerebral cortex, which eventually can lead to cognitive decline. This hypothesis is supported by significant basic science work showing that the combination of Aβ and tau that leads to neurodegeneration, via mechanisms including microglial activation, synaptic spine loss, and suppression of neuronal activity, while either protein in isolation is often compatible with normal functioning [61, 67]. One limitation of this study is that the Aβ PET tracer in HC (18F-Florbetaben) and MCI (18F-Florbetapir) individuals were different. To address this limitation, all the analyses in this paper were replicated with centiloid standard values instead of Aβ SUVR. None of the reported results changed. Since the tau PET tracer was the same in all individuals, in this paper we only reported the analyses with SUVR measures for both pathologies to be more comparable and understandable. Another limitation of this study is that the individuals studied are mostly between the ages of 60 to 70 years old. Considering the aims of this study, this has pros and cons. One of the benefits of this age range is that we were able to detect relatively more individuals with normal levels of Aβ and tau. Conversely, using this age range meant there were a limited number of subjects with abnormal levels of Aβ and tau. We addressed this limitation by adding MCI subjects to our data to detect more abnormal Aβ and tau individuals. A third limitation is that while we have 446 older participants in our analyses, this is still a relatively small sample size when categorizing subjects into four groups based on normal and abnormal levels of Aβ and tau depositions. Therefore, some of our findings did not survive correction for multiple comparisons, especially in groups with a small number of participants. Finally, this study was limited by its use of cross-sectional, rather than longitudinal, data. The relationship between AD pathologies and changes in a sensitive measure like cortical thickness is not easy to analyze through cross-sectional studies. In particular, to provide more compelling evidence with regard to cortical thickening, a longitudinal study with two or more follow-ups is warranted.

## Conclusion

In conclusion, by introducing a new technique based on healthy young brains for categorizing abnormal vs normal Aβ/tau accumulation, we help clarify the effects of Aβ/tau pathologies on neurodegeneration in both populations with normal and abnormal levels of deposition. Our results suggest that, while measuring the normal accumulation of each pathology is critical for understanding the initiation of AD, its pathological progression can be tracked more effectively by measuring the rate of synergy/resonance between Aβ and tau. Altogether, we suggest that considering Aβ’s and tau’s distinct relation with neurodegeneration can only capture the additive individual contribution of each pathology, but the combined (synergetic) effect of these two pathologies has unique toxicity in the brain that should be considered as well. Clarifying these relationships will help establish their role as a potentially critical early biomarker for predicting cognitive decline.

## Data Availability

The data for this project are confidential but may be obtained with Data Use Agreements with the Weill Cornell Medicine and Columbia University Irving Medical Center. Researchers interested in access to the data may contact Dr. Razlighi at. It can take some weeks to negotiate data use agreements and gain access to the data.

## Supplementary material

**Fig. S1:**
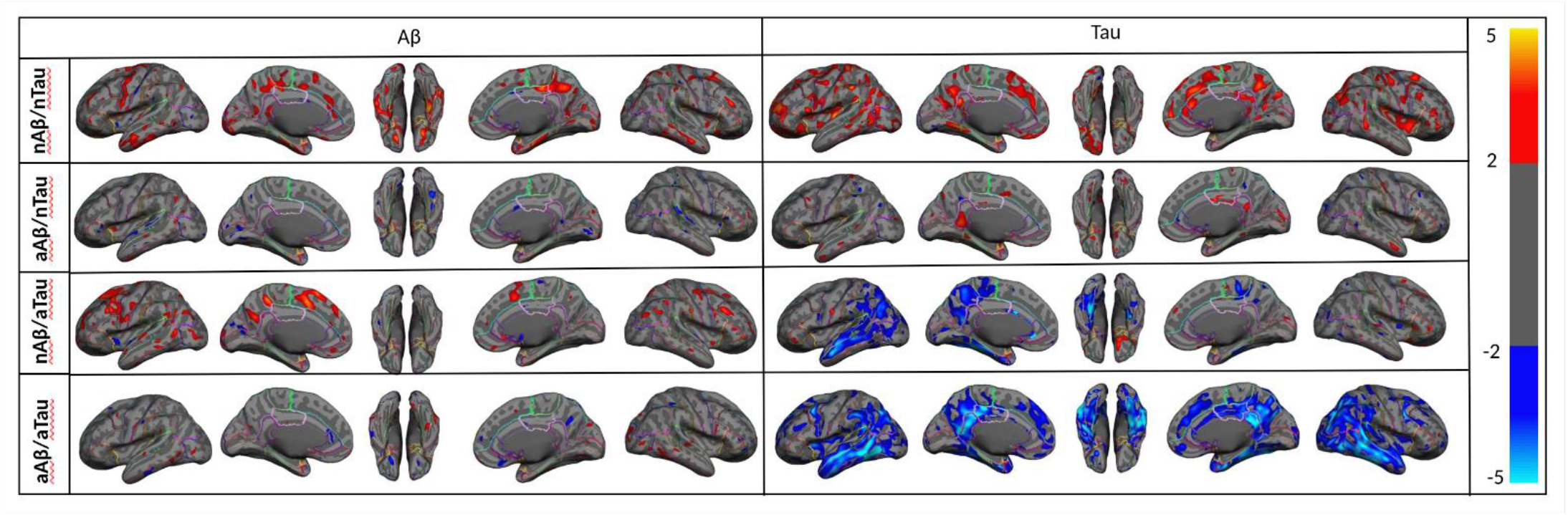
Vertex-wise statistical map (t-value) of association between global Aβ (left column), global tau (right column) pathologies, and cortical thickness throughout the entire cerebral cortex obtained in four categories of participants. (First row, nAβ/nTau; second row, aAβ/nTau; third row, nAβ/aTau; and fourth row, aAβ/aTau). The t-value at each vertex is color-coded with red to yellow colors representing increasing positive t-values and blue to light blue representing decreasing negative t-values and overlaid on the semi-inflated cortical surface of the MNI152 template. The association between global Aβ, global tau pathologies, and cortical thickness was not corrected for multiple comparison corrections.

**Fig. S2:**
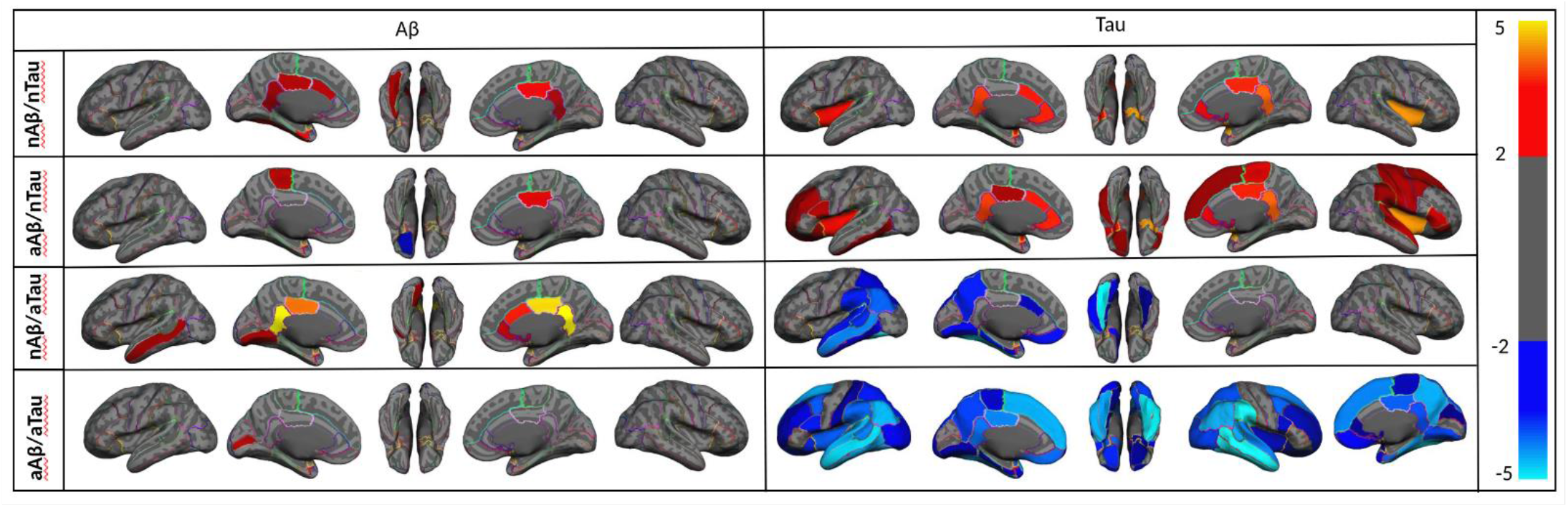
Region-wise statistical map (t-value) of association between regional Aβ (left column), regional tau (right column) pathologies, and regional cortical thickness throughout 68 ROIs obtained in four categories of participants (First row, nAβ/nTau; second row, aAβ/nTau; third row, nAβ/aTau; and fourth row, aAβ/aTau). The t-value at each region is color-coded with red to yellow colors representing increasing positive t-values and blue to light blue representing decreasing negative t-values and overlaid on the semi-inflated cortical surface of the MNI152 template. Association between regional Aβ, tau pathologies, and cortical thickness was not corrected for family-wise errors.

**Fig. S3:**
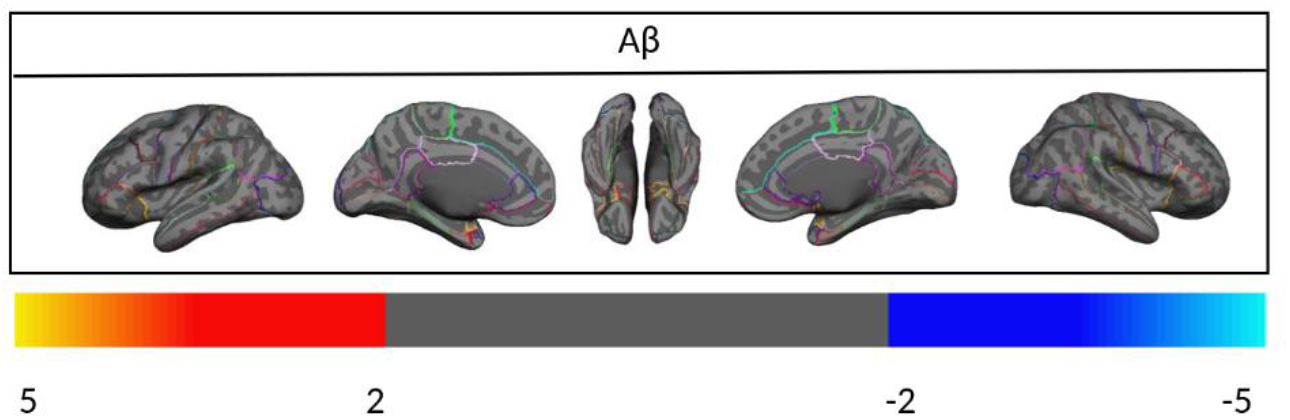
Vertex-wise statistical map (t-value) of association between global Aβ pathology and cortical thickness throughout the entire cerebral cortex obtained in young subjects. The t-value at each vertex is color-coded with red to yellow colors representing increasing positive t-values and blue to light blue representing decreasing negative t-values and overlaid on the semi-inflated cortical surface of the MNI152 template. The association between global Aβ pathology and cortical thickness was not corrected for multiple comparison correction.

**Fig. S4:**
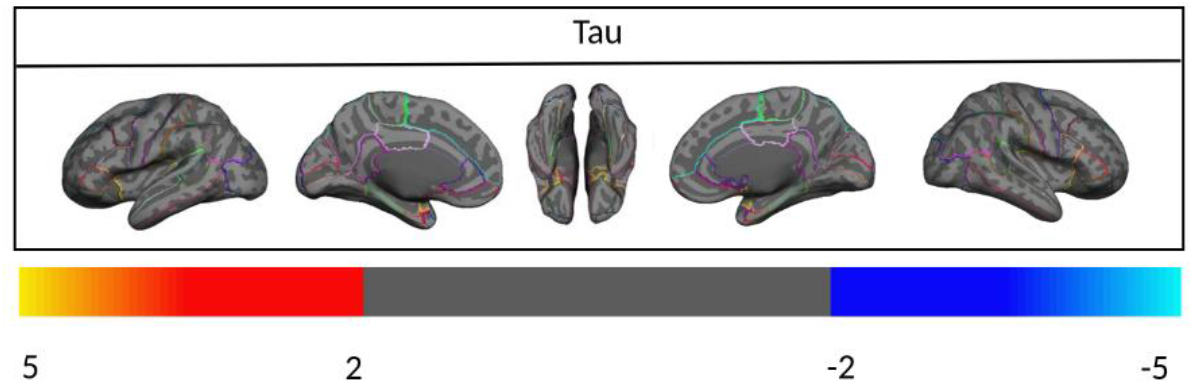
Vertex-wise statistical map (t-value) of association between global tau pathology and cortical thickness throughout the entire cerebral cortex obtained in young subjects. The t-value at each vertex is color-coded with red to yellow colors representing increasing positive t-values and blue to light blue representing decreasing negative t-values and overlaid on the semi-inflated cortical surface of the MNI152 template. Association between global tau pathology and cortical thickness was not corrected for multiple comparison correction.

**Fig. S5:**
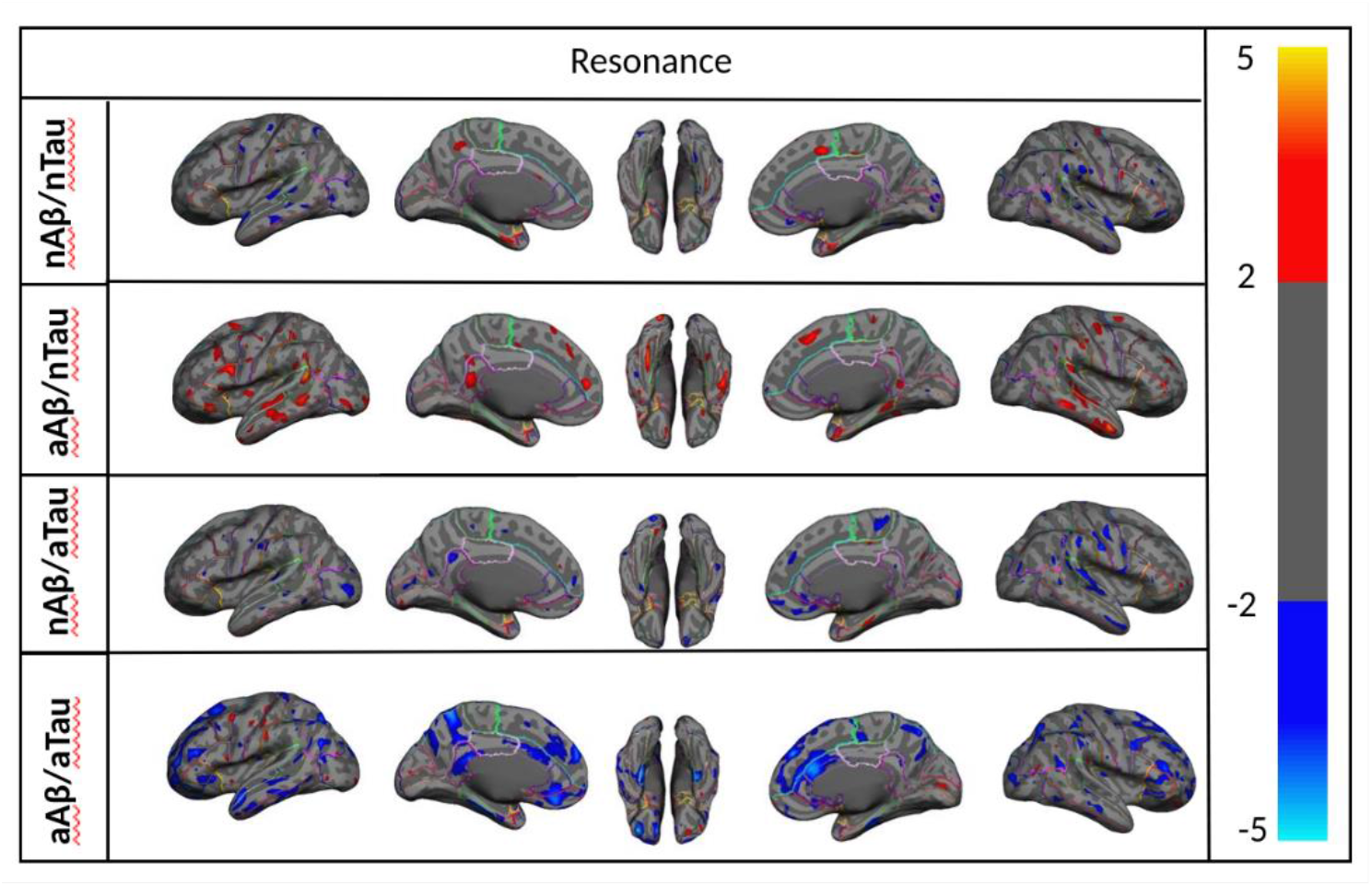
Vertex-wise statistical map (t-value) of the association between resonance and cortical thickness throughout the entire cerebral cortex obtained in four categories of participants in the aAβ/aTau group. The t-value at each vertex is color-coded with a heatmap where red to yellow colors represents increasing positive t-values and blue to light blue represents decreasing negative t-values overlaid on the semi-inflated cortical surface of the MNI152 template. Associations between resonance and cortical thickness were not corrected for multiple comparison correction.

**Fig. S6:**
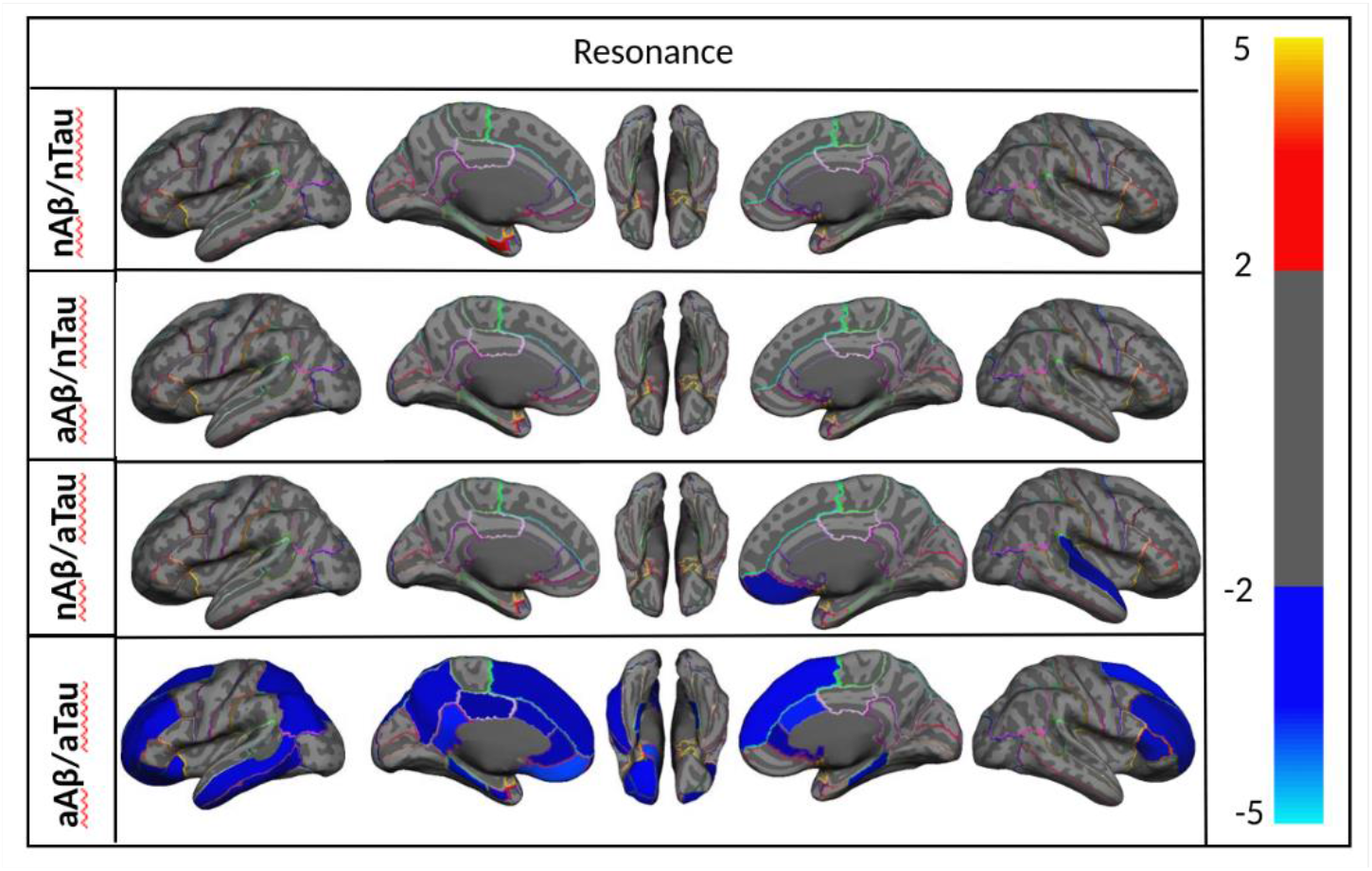
(a) Region-wise statistical map (t-value) of the association between resonance and cortical thickness throughout 68 ROIs obtained in the aAβ/aTau group. The t-value at each region is color-coded with a heatmap where red to yellow colors represents increasing positive t-values and blue to light blue represents decreasing negative t-values overlaid on the semi-inflated cortical surface of the MNI152 template. Association between resonance and cortical thickness was not corrected for family-wise errors.

